# Measured intrapatient radiomic variability as a predictor of treatment response in multi-metastatic soft tissue sarcoma patients

**DOI:** 10.1101/2025.04.11.25325700

**Authors:** Caryn Geady, James J Bannon, Shagheyegh Reza, Laura Madanat-Harjuoja, Denise Reinke, Scott Schuetze, Brian Crompton, Andrew Hope, Benjamin Haibe-Kains

## Abstract

Radiomics offers a non-invasive approach to tumor characterization, yet its application in metastatic cancers is limited by intertumor heterogeneity—variability in radiomic phenotypes across lesions within the same patient. We introduce Measured Intrapatient Radiomic Variability (MIRV), a novel metric quantifying heterogeneity using standard-of-care imaging. Applied to 397 metastatic soft-tissue sarcoma (STS) patients from the SARC021 trial, MIRV was calculated from pretreatment CT scans using pairwise Euclidean distance and cosine dissimilarity between lesions. Euclidean distance captures absolute differences in radiomic features, while cosine dissimilarity assesses variation in feature patterns independent of magnitude.

Higher MIRV correlated with greater variability in tumor-specific response classification (TSRC) and volumetric response, independent of baseline tumor volume. In a subset with liquid biopsy data, MIRV showed a moderate association with ctDNA positivity, suggesting links to molecular heterogeneity. While MIRV was not prognostic for overall survival (OS) in the full cohort, higher MIRV was significantly associated with worse survival in leiomyosarcoma patients (n=165, p=0.007). These findings establish MIRV as a biomarker for intertumor heterogeneity, with potential to predict mixed treatment responses and guide personalized therapy in metastatic STS. Future studies should assess its relevance across other tumor types and therapeutic settings.

## Introduction

Radiological imaging and its quantitative analysis, known as radiomics, is a rapidly evolving field with significant promise for precision oncology. By enabling non-invasive characterization of tumor phenotypes [1], radiomics has advanced our understanding of intratumoral spatial heterogeneity in localized cancers, where variations in tumor biology and resistance mechanisms have been well documented [1–3]. However, in the metastatic setting, the focus shifts to intertumor heterogeneity— the variability in radiomic phenotypes across multiple lesions within the same patient. This heterogeneity is clinically relevant, as it may influence disease progression, therapeutic response, and the success of personalized treatment strategies [4–6].

Despite growing recognition of intertumor heterogeneity [3,4], radiomic analyses in this context often depend on specific signals or modalities that are not universally available, limiting their broader applicability. To overcome this limitation, we introduce measured intrapatient radiomic variability (MIRV), a novel metric designed to quantify intertumor heterogeneity using standard-of-care imaging. By leveraging widely available imaging data, MIRV provides a scalable and clinically applicable framework for characterizing tumor heterogeneity without the need for additional imaging acquisition beyond routine clinical practice.

In this study, we apply MIRV to metastatic soft-tissue sarcomas (STS) as a representative cancer type to investigate its biological and clinical implications. STS represents an ideal setting for studying tumor heterogeneity due to its extensive variability across clinical, histological, molecular, and radiological dimensions [7,8]. We hypothesize that greater differences in radiomic phenotypes between metastatic lesions are associated with variability in treatment response, potentially impacting personalized therapeutic decision-making. By evaluating MIRV as a biomarker for early treatment response, we demonstrate the potential of MIRV in guiding timely clinical interventions and improving patient outcomes in metastatic cancer.

Our analysis reveals that MIRV captures clinically meaningful variability in treatment response. Higher MIRV was associated with increased heterogeneity in volumetric response, independent of baseline tumor volume. In a subset of patients with liquid biopsy data, MIRV correlated with ctDNA positivity, suggesting a potential link to greater release of tumor DNA into the circulatory system, which may reflect higher tumor necrosis, cell turnover, or hypoxia. While MIRV was not prognostic for overall survival in the full cohort, it was significantly associated with worse survival in leiomyosarcoma patients, highlighting its potential subtype-specific relevance. These findings establish MIRV as a novel biomarker for intertumor heterogeneity, with implications for predicting mixed treatment responses and refining personalized therapeutic strategies in metastatic STS.

## Results

This study introduces Measured Intrapatient Radiomic Variability (MIRV) as a novel metric to quantify intertumor heterogeneity using standard-of-care imaging. We applied MIRV to a cohort of multi-metastatic soft-tissue sarcoma (STS) patients, extracting and reducing radiomic features from pretreatment CT scans. We then assessed MIRV’s association with treatment response metrics— including volumetric response, tumor-specific response classification (TSRC), and ctDNA positivity— and evaluated its prognostic relevance through statistical analyses (**Figure 1**).

**Figure 1.**
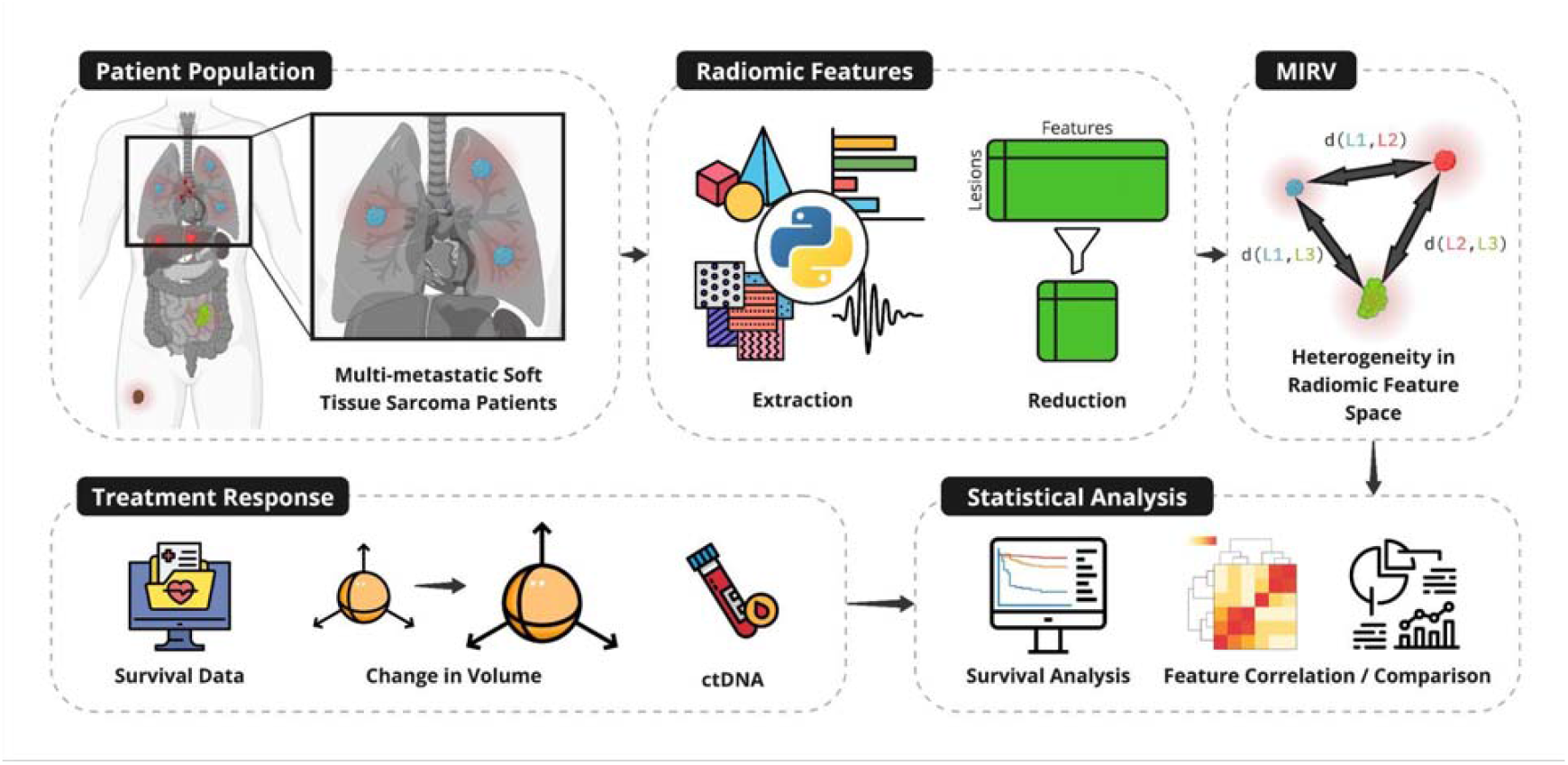
Evaluating intertumor heterogeneity using Measured Intrapatient Radiomic Variability (MIRV) as a biomarker of treatment response and clinical covariates. Multi-metastatic soft tissue sarcoma patients were analyzed (top left). Radiomic features were extracted from baseline imaging and reduced to capture non-redundant tumor-specific characteristics (top center). Intertumor heterogeneity was quantified using MIRV, defined by distances between tumors in radiomic feature space (top right). Treatment response metrics, including survival data, tumor-specific volumetric changes, and ctDNA positivity, were collected (bottom left). Statistical analyses were performed to correlate MIRV with treatment outcomes, including survival analysis and feature comparison (bottom right).

### Patient Cohort and Data Collection

We retrospectively analyzed a cohort of metastatic soft-tissue sarcoma (STS) patients from SARC021, a phase III clinical trial (TH-CR-406/SARC021, NCT01440088) conducted by the Sarcoma Alliance for Research through Collaboration (SARC) [9]. The study included patients with locally advanced, unresectable, or metastatic disease. To assess mixed intrapatient response variability (MIRV), we focused on patients with multiple metastases contoured on pre-treatment computed tomography (CT) scans. Data collection was approved by institutional ethics review (REB #20-5707), and full trial details can be found in [9].

The dataset comprised 397 patients, including individuals diagnosed with leiomyosarcoma (41%), liposarcoma (14%), undifferentiated pleomorphic sarcoma (12%), and other STS subtypes (33%). For all patients, we collected pretreatment CT scans and extracted radiomic features alongside clinical characteristics such as RECIST 1.1 assessments and histological subtypes. A separate study, independent of SARC021, assessed the feasibility of detecting circulating tumor DNA (ctDNA) in a subset of the leiomyosarcoma patients [10], which we were able to incorporate into our analysis. For this subset of patients, ctDNA status was recorded as either positive or negative based on the presence or absence of detectable ctDNA levels, assessed using ichorCNA technology [10]. Additionally, for patients with pulmonary metastases, we examined post-treatment CT scans to evaluate volumetric response. Tumor-specific response classification (TSRC) was applied using volumetric measurements for the lung subset, while RECIST-based response was assessed for the full cohort.

### MIRV and Response Metrics

Radiomic feature extraction was performed on CT images using PyRadiomics (version 3.0.1) [11]. To reduce redundancy, a stepwise feature reduction process was applied, removing low-variance features, those correlated with tumor volume (|ρ| > 0.1), and highly correlated features (|ρ| > 0.7) [12]. The remaining features were used to compute MIRV metrics (**Supplementary Tables 1 and 2**). MIRV was then quantified using pairwise cosine dissimilarity and Euclidean distance between tumor radiomic feature vectors within each patient. Cosine dissimilarity (1 − cosine similarity) captures differences in feature orientation [13], reflecting whether tumors share similar radiomic patterns regardless of absolute feature values. Euclidean distance measures the overall difference between feature vectors, incorporating both magnitude and direction [13]. Together, these metrics provide a comprehensive assessment of intertumor heterogeneity.

**Table 1.**
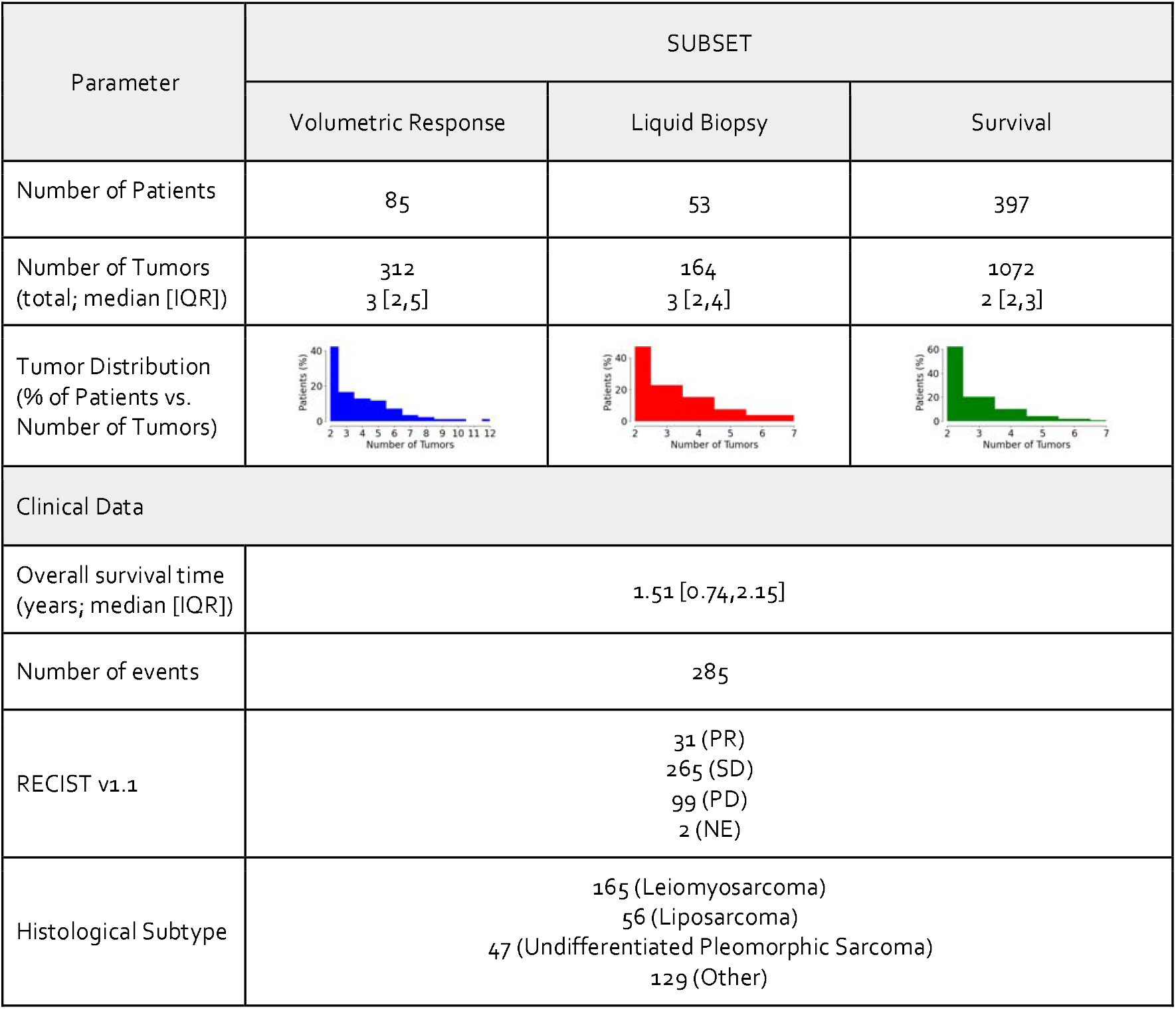
Demographics and clinical information for the dataset used in this study. Statistics surrounding the number of tumors per patient and survival are expressed in terms of the median and the interquartile range (IQR).

**Table 2.**
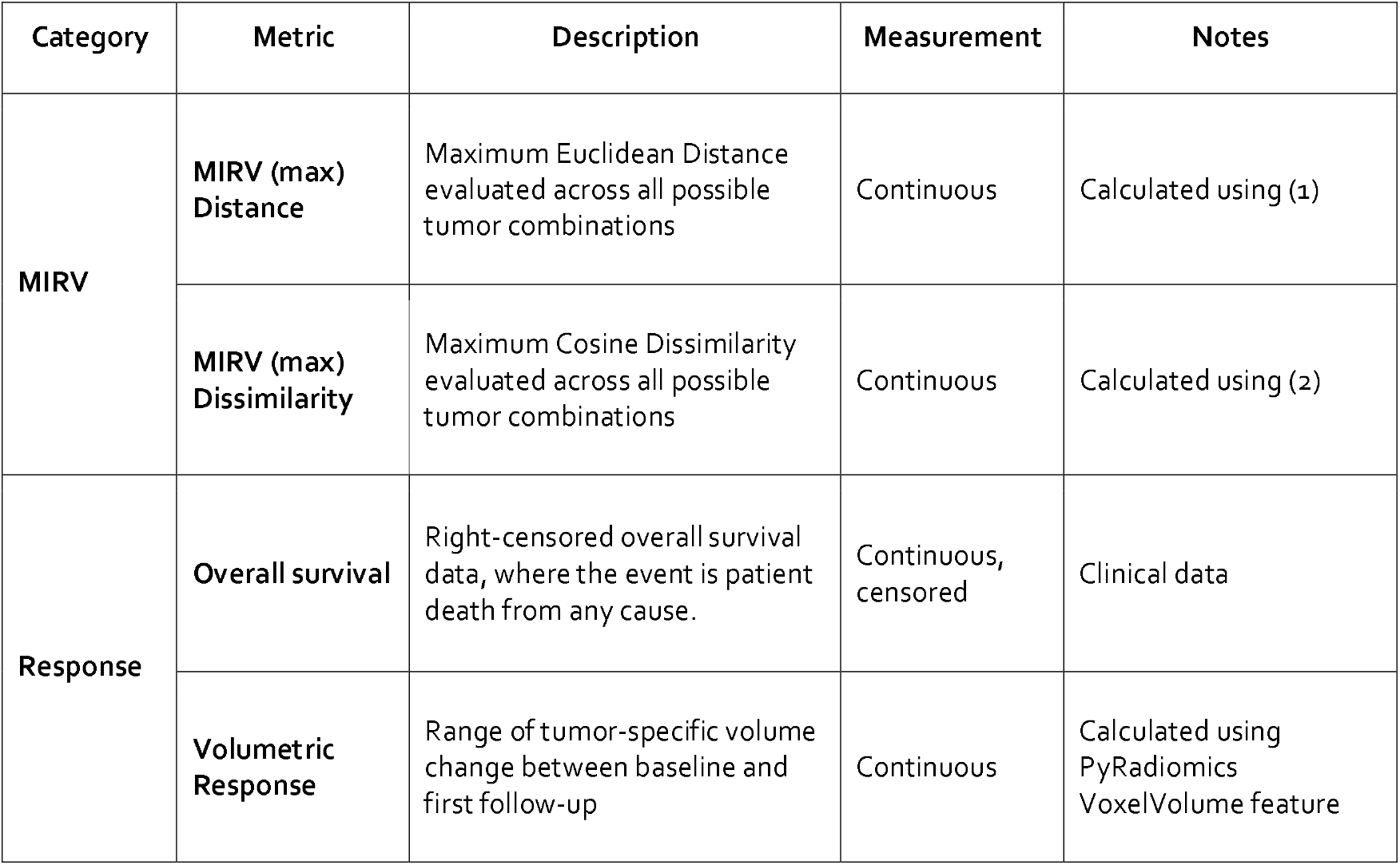
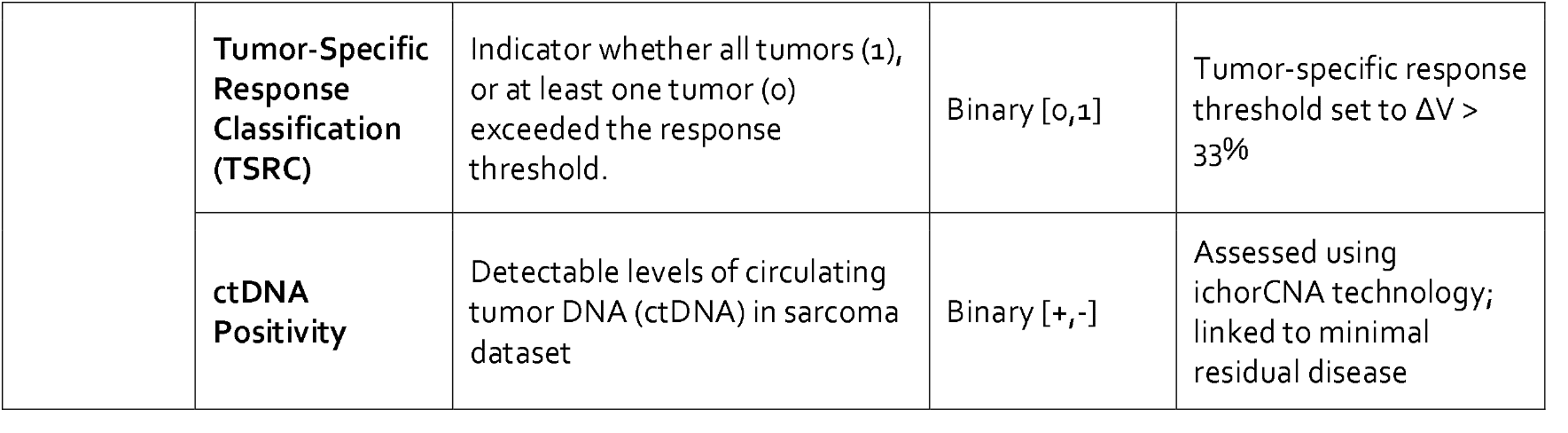
Summary of MIRV and response metrics, including overall survival, volumetric response, tumor-specific response classification (TSRC) and liquid biopsy data.

We evaluated the relationship between MIRV and overall survival (OS), volumetric response, tumor-specific response classification (TSRC), and ctDNA positivity (**Table 2**). Survival serves as a definitive clinical endpoint, capturing long-term patient outcomes; however, it can be influenced by undocumented factors, making attribution to tumor heterogeneity challenging. Volumetric response measures differences in tumor volume change over time, providing an objective assessment of treatment effects across lesions. While sensitive to response heterogeneity, it may not fully capture non-size-based treatment effects. TSRC categorizes whether all tumors in a patient met a predefined 33% volume reduction threshold [14], offering a simplified binary measure of response consistency. However, it does not account for partial responses or variations in response magnitude. Finally, ctDNA positivity from liquid biopsy analysis may serve as a non-invasive indicator of tumor burden and tumor response to therapy. While valuable, ctDNA detection may be influenced by tumor shedding dynamics and assay sensitivity [15,16]. Additional details are provided in the Methods section.

### Association Between MIRV, Tumor Response, and Liquid Biopsy Markers

Our analysis revealed significant associations between MIRV, TSRC, volumetric response, and ctDNA positivity (**Figure 2**). Both MIRV metrics exhibited a strong negative correlation with Complete Tumor Response, suggesting that greater intertumor heterogeneity may be linked to non-uniform tumor responses within individual patients. Additionally, MIRV demonstrated a moderate correlation with volumetric response, independent of baseline tumor volume, reinforcing that radiomic heterogeneity captures treatment-related changes beyond tumor burden alone. This distinction is critical, as prior studies have shown that radiomic features can act as surrogates for volume when predicting survival [12,17,18]. By implementing three baseline volume measures as controls, we confirmed that baseline tumor volume does not inherently predict volumetric response, underscoring MIRV’s role in reflecting biologically meaningful variability in treatment outcomes.

**Figure 2.**
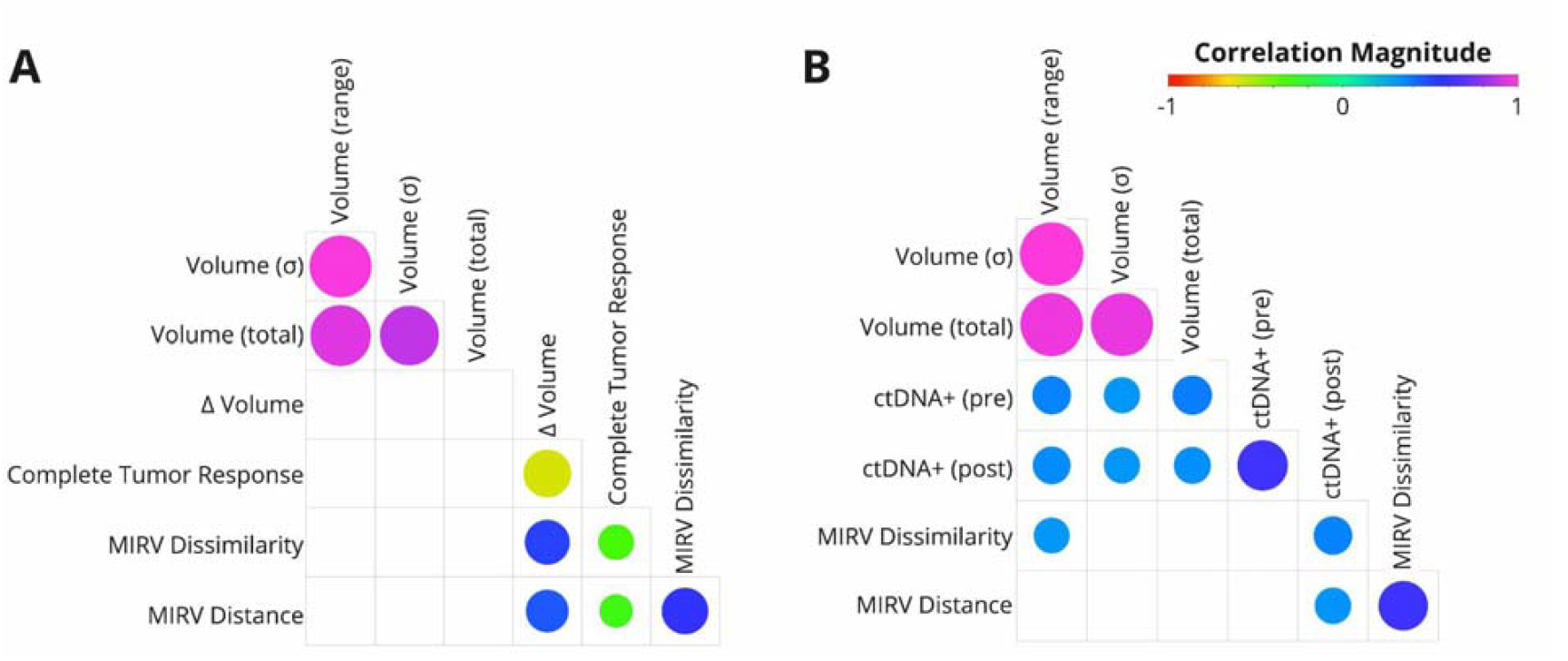
Correlation between MIRV, tumor response, and ctDNA positivity. (A) Relationships between MIRV, TSRC (Complete Tumor Response), and volumetric changes in the volumetric response subset. (B) Relationships between MIRV, and ctDNA pre- and post-treatment in the liquid biopsy subset. Baseline tumor volume characteristics were included as a control. Circle size represents correlation strength (FDR), with colors indicating Spearman’s correlation coefficients (ρ) according to the scale bar. In cases where the association was not significant (FDR > 0.05), circles are not displayed.

In the liquid biopsy subset (**Figure 2B**), MIRV metrics showed a moderate correlation with post-treatment ctDNA positivity. Given that ctDNA reflects tumor burden [19,20], its association with baseline volume metrics was expected. However, MIRV Distance remained largely independent of baseline tumor volume, reinforcing that intertumor heterogeneity captures intrinsic tumor biology beyond size alone. The differing correlation patterns observed between MIRV (cosine vs. Euclidean distance) and baseline volume measures suggest that these metrics may capture complementary aspects of radiomic heterogeneity, with cosine distance being more sensitive to differences in feature composition rather than absolute magnitude. These findings highlight MIRV’s potential as a biomarker linking radiological and molecular indicators of treatment response.

### MIRV Exhibits Context-Specific Survival Implications

We used a multivariable Cox proportional hazards model to assess the prognostic significance of MIRV while controlling for known clinical factors, namely histologic classification, observed baseline volume, performance status, patient age and RECIST classification. This approach allowed us to determine whether MIRV provided independent prognostic information beyond established clinical variables. After adjusting for these factors, no significant association between MIRV and overall survival was observed in the full cohort. However, subgroup analysis was conducted based on a significant interaction term between MIRV and histological subtype identified in the multivariable model (**Figure 3A**). In these subgroup analyses, log-rank tests revealed that higher MIRV was associated with significantly worse overall survival in the leiomyosarcoma cohort (n=165, log-rank p = 0.007 for MIRV Dissimilarity (**Figure 3B**) and p = 0.06 for MIRV Distance). These findings suggest that, while MIRV does not have prognostic significance across the full cohort, it may serve as a relevant prognostic marker in specific patient subgroups.

**Figure 3.**
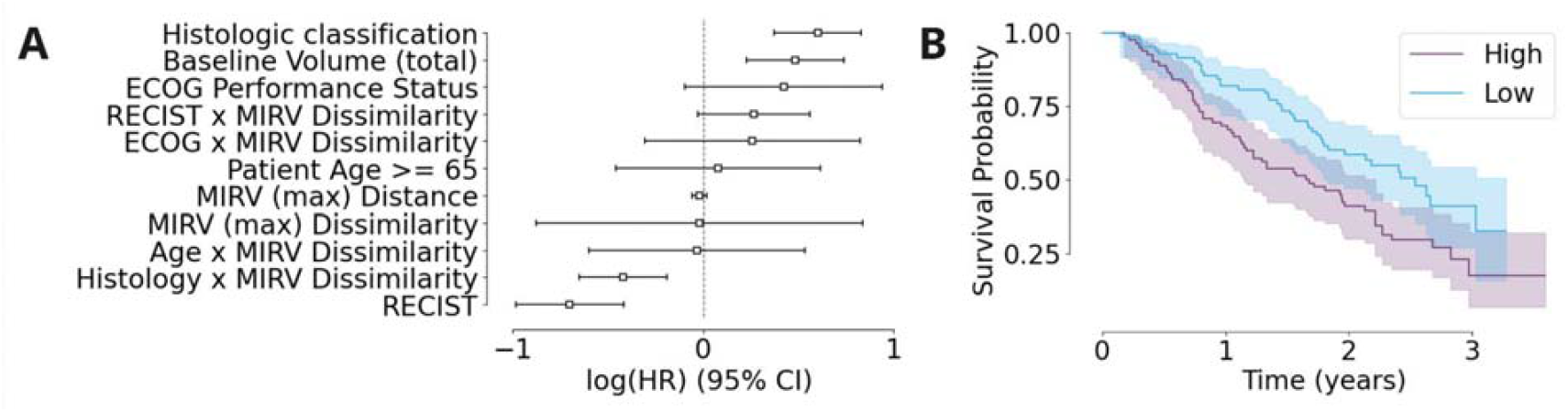
(A) Forest plot showing the log(HR) and 95% confidence intervals for multiple clinical and imaging-based covariates, including MIRV (max) Dissimilarity and Distance. (B) Kaplan-Meier survival curves stratified by MIRV Dissimilarity, with high MIRV (blue) associated with worse survival probability compared to low MIRV (orange). Shaded areas represent the 95% confidence intervals.

## Discussion

Our study introduces MIRV as a novel imaging-based biomarker for assessing tumor heterogeneity and treatment response. While MIRV was not significantly associated with survival across the entire cohort, sarcomas are highly heterogeneous. In histologic subgroup analysis, MIRV showed potential prognostic relevance, particularly in leiomyosarcoma. Notably, MIRV exhibited complementary associations with clinically relevant response metrics, including TSRC, tumor volume changes, and ctDNA positivity, highlighting its ability to capture distinct aspects of treatment response. Given the complexity of survival as an outcome measure, these findings suggest that MIRV provides independent and biologically meaningful insights into tumor heterogeneity that may inform prognosis.

Traditional radiomic features have been proposed as descriptors of tumor heterogeneity, but many primarily act as surrogates for baseline tumor volume [12,18]. Our approach extends radiomics beyond the conventional framework—where typically only one tumor per patient is analyzed [21] —to a paradigm that incorporates multiple tumors per patient. This enables a more comprehensive assessment of tumor heterogeneity and its relationship to treatment response, independent of baseline tumor burden. Furthermore, MIRV is evaluated on a per-patient basis, mitigating image-related biases common in multi-institutional studies caused by variations in imaging protocols and equipment [22]. This normalization enhances MIRV’s applicability across diverse patient populations.

The clinical implications of our findings highlight MIRV’s potential to supplement existing treatment response metrics, offering a new dimension for oncological decision-making. To further validate its utility, prospective studies in other cancer types and investigations into its predictive power for targeted therapies, such as immunotherapy and radiation therapy, are warranted. Despite these promising findings, our study has limitations. The retrospective design and dataset-specific biases may limit generalizability, and additional research is needed to optimize MIRV thresholds for clinical use. A key challenge is the scarcity of datasets containing sequential volumetric segmentations, essential for tracking tumor kinetics over time. However, our MIRV analysis pipeline is openly available, and we encourage researchers to apply it to their own datasets or collaborate on future research to expand its clinical relevance.

## Conclusion

Our study demonstrates that MIRV provides unique insights into tumor biology, heterogeneity, and treatment response, beyond traditional prognostic markers. By capturing tumor-specific variability, MIRV may enhance patient stratification and guide more personalized therapeutic strategies. Integrating MIRV into clinical workflows could refine response assessment and improve precision oncology. While MIRV’s direct prognostic utility remains to be fully established, its role in characterizing treatment-related heterogeneity suggests a potential avenue for further research and clinical translation.

## Methods

### Data Processing and Analysis

#### Radiomic Feature Handling

Radiomic feature extraction was performed on CT images using PyRadiomics (version 3.0.1) [11]. To address feature redundancy and improve downstream analyses, a stepwise feature reduction pipeline was implemented. First, features with low variance were removed using a variance filter (low variance was defined as less than the median variance across all features). Next, features demonstrating an absolute Spearman rho correlation with tumor volume greater than 0.1 were excluded [12]. Finally, any remaining features with an absolute Spearman correlation exceeding 0.7 with any other feature were discarded. The resulting feature set was utilized to calculate MIRV.

#### MIRV Metrics

To calculate MIRV metrics, we developed a function to compute pairwise cosine dissimilarity (1−cosine similarity) and Euclidean distance between tumors for each patient. Cosine dissimilarity capture differences in feature orientation, making it robust to variations in absolute feature magnitudes, while Euclidean distance reflects overall dissimilarity in feature space, accounting for both magnitude and direction. Together, these metrics provide complementary insights into intertumor heterogeneity— cosine dissimilarity emphasizing shape-based differences and Euclidean distance capturing absolute divergence in tumor characteristics. For each patient, pairwise metrics were calculated across all possible tumor combinations, and the maximum values were used to summarize intertumor relationships, highlighting the most divergent tumor pair per patient ((1),(2)).

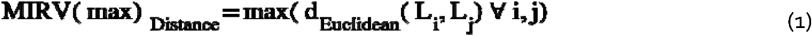

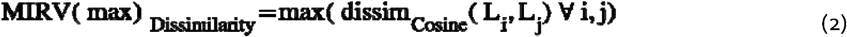

#### Response Metrics

Response metrics included overall survival (OS) expressed in years, and volumetric response assessed at a post-treatment timepoint, tumor-specific response classification and liquid biopsy data (Table 2).

#### Volumetric Response

Volumetric response was assessed by calculating the relative change in tumor volume between baseline and first follow-up scans. Contours delineated at both time points were used to perform these calculations. The PyRadiomics **‘VoxelVolume’** feature served as the measure of tumor volume at baseline and follow-up. Relative volume change was expressed as a percentage. At the patient level, the range in volume change across all tumors was assessed and reported as a percentage.

#### Tumor-Specific Response Classification (TSRC)

To assess intertumor variability in treatment response, patients were categorized based on the concordance of volumetric changes across their metastatic tumors. Tumor responses were evaluated using a predefined volume change threshold (ΔV > 33%) [14]. Patients were classified as having Complete Tumor Response if all tumors exhibited a change in volume that did not exceed the threshold. Patients that had at least one tumor that exceeded the response threshold, including cases where all tumors exceeded the threshold, were classified as Non-/Partial Tumor Response.

#### Liquid Biopsy Data

For this dataset, we use the presence of detectable ctDNA levels as a metric for therapy response, which was recorded as binary data (positive or negative) based on the presence or absence of detectable levels of ctDNA in the sample. The collection and processing of ctDNA data for the sarcoma dataset are described in [10]; briefly, a positive ctDNA test indicates microscopic amounts of cancer cells, often referred to as minimal residual disease (MRD), in the patient’s blood. The levels of detectable ctDNA were assessed using the ichorCNA technology.

### Statistical Analysis

We conducted two main types of analyses: (1) correlation between MIRV and treatment response metrics, and (2) survival analysis evaluating MIRV alongside clinical and radiological metrics.

#### Correlation Between MIRV and Treatment Response

Spearman’s rank correlation coefficient was used to evaluate associations between MIRV and treatment response metrics. For the lung metastasis subset, MIRV was correlated with changes in tumor volume and TSRC. For the liquid biopsy subset, MIRV was compared with ctDNA positivity to investigate its relationship with molecular response. Given the potential sensitivity of MIRV to baseline tumor volume, we also considered three baseline volumetric measures: (1) range in tumor volumes (absolute difference between the smallest and largest tumor per patient), (2) standard deviation of tumor volumes, and (3) total tumor volume. Baseline volume metrics were derived using the PyRadiomics VoxelVolume feature, consistent with volumetric response analyses. Multiple testing correction was applied using the False Discovery Rate (FDR) method in the statsmodels package.

#### Survival Analysis

The prognostic value of MIRV was first assessed using continuous MIRV values in a Cox proportional hazards model to evaluate its association with overall survival (OS). Additionally, we computed the Hazard Ratio to quantify the predictive ability of MIRV relative to clinical covariates, including performance status, patient age, histological subtype, RECIST response, and total tumor burden (measured by total volume). Subgroup analyses were conducted based on interaction terms identified in the multivariable model, with Kaplan-Meier survival analysis and log-rank tests performed to evaluate overall survival differences within specific patient subgroups. All survival analyses were conducted using the lifelines package in Python.

## Data Availability and Research Reproducibility

Imaging and clinical data for the SARC021 trial are not publicly available but can be obtained through SARC with a DUA (Contact for details on using data from SARC). The de-identified radiomic features and clinical information, along with the code for data cleaning and analysis that underlie the results reported in the article are publicly available. The computer code is available from https://github.com/bhklab/MIRV, with relevant parameter files to replicate the software environment used to generate the results in this paper. Notably, the environment for feature extraction and analysis was configured using Pixi, enabling compatibility across Windows, Mac, and Linux operating systems. Additionally, the full software environment, processed data and computer code are available from https://codeocean.com/capsule/8750010/tree/v1.

## Acknowledgements

The authors would like to thank the Sarcoma Alliance for Research through Collaboration (SARC) for access to the imaging data used in this study.

## Author contributions

CG was responsible for conceptualization, methodology, formal analysis, investigation, data curation, writing the original draft, and visualization. SR and JB contributed to investigation and statistical rigor. DR and SS contributed domain-specific insights towards interpretation of findings. BHK and AH contributed to conceptualization, data and computational resources, supervision, and funding acquisition. All authors reviewed the manuscript.

## Additional Information

The authors declare that they have no competing interests. Research reported in this publication was supported by the National Cancer Institute of the National Institutes of Health under Award Number P50CA272170. The content is solely the responsibility of the authors and does not necessarily represent the official views of the National Institutes of Health.

## Supplemental

**Table S1.** Summary of radiomic features used to calculate MIRV for each subset of the patient data: volumetric response, liquid biopsy and survival. Numbers of features used for the calculation are recorded at different stages of the unsupervised feature reduction step.

| Number of Features | SUBSET |  |  |
| --- | --- | --- | --- |
|  | Volumetric Response | Liquid Biopsy | Survival |
| Total Extracted | 1218 |  |  |
| High Variance, Uncorrelated to Volume | 248 | 265 | 261 |
| High Variance, Uncorrelated to Volume and Other Features | 16 | 10 | 13 |

**Table S2.** Summary of radiomic features used to calculate MIRV for each subset of the patient data: volumetric response, liquid biopsy and survival. Five features are used in all three subsets, and sixteen features are used by at least two subsets. Five features are unique to the volumetric response subset and three features are unique to the liquid biopsy subset.

| Radiomics Features |  |  | SUBSET |  |  |
| --- | --- | --- | --- | --- | --- |
| Filter | Class | Name | Volumetric Response | Liquid Biopsy | Survival |
| Gradient | GLSZM | SmallAreaEmphasis | ? | ? | ? |
| Wavelet-HHL | GLCM | ClusterShade | ? | ? | ? |
| Wavelet-HHL | FirstOrder | Median | ? |  | ? |
| Wavelet-HHH | FirstOrder | Median | ? |  | ? |
| SquareRoot | GLCM | ClusterShade | ? |  | ? |
| Wavelet-HLH | FirstOrder | Median | ? |  | ? |
| Wavelet-LHH | FirstOrder | Median | ? |  | ? |
| Wavelet-LLH | GLCM | ClusterShade | ? |  | ? |
| Exponential | GLSZM | SmallAreaEmphasis |  | ? | ? |
| Wavelet-HLH | GLSZM | SmallAreaEmphasis |  | ? | ? |
| Wavelet-LHL | GLCM | ClusterShade |  | ? | ? |
| Wavelet-LLL | GLCM | Correlation |  | ? | ? |
| Wavelet-HLH | GLCM | ClusterShade | ? | ? |  |
| Wavelet-HLL | GLCM | ClusterShade | ? | ? |  |
| Wavelet-LHH | GLCM | ClusterShade | ? | ? |  |
| Wavelet-HLL | FirstOrder | Kurtosis |  | ? |  |
| Wavelet-LLH | GLCM | Correlation | ? |  |  |
| Wavelet-HHH | GLSZM | SmallAreaEmphasis | ? |  |  |
| Square | GLSZM | SmallAreaEmphasis | ? |  |  |
| Logarithm | FirstOrder | InterquartileRange | ? |  |  |
| Logarithm | GLDM | LargeDependenceHighGrayLevelEmphasis | ? |  |  |
| Wavelet-LHH | GLSZM | SmallAreaEmphasis |  |  | ? |

**Table S3.** Hazard ratios (HR) with 95% confidence intervals (CI), z-scores, and p-values for key clinical and radiomic variables in a multivariable Cox proportional hazards model assessing overall survival in metastatic soft-tissue sarcoma patients. Significant predictors of worse survival include histologic classification (HR = 1.82, p < 0.005), and baseline tumor volume (HR = 1.57, p < 0.005). RECIST response was significantly associated with better survival (HR = 0.54, p < 0.005). MIRV-based metrics (dissimilarity and distance) were not significant prognostic factors (p = 0.58 and p = 0.94, respectively), but the histology-MIRV interaction term (Dissimilarity) was significantly associated with better survival (HR = 0.68, p < 0.005).

| Variable | Hazard Ratio [95% conf.int.] | z | p |
| --- | --- | --- | --- |
| Patient Age $\geq 65$ | 0.88 [0.50,1.56] | -0.43 | 0.67 |
| ECOG Performance Status | 1.46 [0.84,2.53] | 1.35 | 0.18 |
| Histologic classification | 1.82 [1.44,2.31] | 4.94 | <0.005 |
| MIRV (max) Dissimilarity | 1.21 [0.62,2.38] | 0.55 | 0.58 |
| MIRV (max) Distance | 1.00 [0.96,1.05] | 0.07 | 0.94 |
| Baseline Volume (total) | 1.57 [1.21,2.05] | 3.35 | <0.005 |
| RECIST | 0.54 [0.40,0.72] | -4.08 | <0.005 |
| ECOG x MIRV Dissimilarity | 1.26 [0.68,2.37] | 0.73 | 0.46 |
| Histology x MIRV Dissimilarity | 0.68 [0.52,0.87] | -3.04 | <0.005 |
| Age x MIRV Dissimilarity | 1.18 [0.63,2.21] | 0.51 | 0.61 |
| RECIST x MIRV Dissimilarity | 1.14 [0.82,1.59] | 0.79 | 0.43 |

